# Psychosocial factors and hospitalisations for COVID-19: Prospective cohort study of the general population

**DOI:** 10.1101/2020.05.29.20100735

**Authors:** G. David Batty, Ian J. Deary, Michelle Luciano, Drew M. Altschul, Mika Kivimäki, Catharine R. Gale

**Author notes:** Correspondence: David Batty, Department of Epidemiology and Public Health, University College London, 1-19 Torrington Place, London, UK, WC1E 6BT. E.

## Abstract

**Objective:** To examine the association of a range of psychosocial factors with hospitalisation for COVID-19.

**Design:** Prospective cohort study.

**Setting:** England.

**Participants:** UK Biobank comprises around half a million people who were aged 40 to 69 years at study induction between 2006 and 2010 when information on psychosocial factors and covariates were captured.

**Main outcome measure:** Hospitalisation for COVID-19 in England between 16^th^ March and 26^th^ April 2020 as provided by Public Health England.

**Results:** There were 908 hospitalisations for COVID-19 in an analytical sample of 431,051 people. In age- and sex-adjusted analyses, an elevated risk of COVID-19 was related to disadvantaged levels of education (odds ratio; 95% confidence interval: 2.05; 1.70, 2.47), income (2.00; 1.63, 2,47), area deprivation (2.20; 1.86, 2.59), occupation (1.39; 1.14, 1.69), psychological distress (1.58; 1.32, 1.89), mental health (1.50; 1.25, 1.79), neuroticism (1.19; 1.00, 1.42), and performance on two tests of cognitive function – verbal and numerical reasoning (2.66; 2.06, 3.34) and reaction speed (1.27; 1.08, 1.51). These associations were graded (p-value for trend ≤0.038) such that effects were apparent across the full psychosocial continua. After mutual adjustment for these characteristics plus ethnicity, comorbidity, and lifestyle factors, only the relationship between lower cognitive function as measured using the reasoning test and a doubling in the risk of the infection remained (1.98; 1.38, 2.85).

**Conclusion:** A range of psychosocial factors revealed associations with hospitalisations for COVID-19 of which the relation with cognitive function was most robust to statistical adjustment.

**Box:**

What is already known on this subject
- Given the recent discovery of COVID-19, its risk factors are not well understood.
- The little evidence that is available has been gleaned from prognostic studies of disease progression and death.
- We are not aware of any studies examining the role of psychosocial factors in the prevention of serious cases of the infection.

What this study adds
- A higher risk of hospitalisation for COVID-19 was evident at disadvantaged levels of education, income, area deprivation, occupation, mental health, neuroticism, and cognitive function.
- After taking into account multiple confounding factors, the strongest association was apparent for cognitive function, a potential marker of health literacy.

## Introduction

With outbreaks by then reported across 114 countries, the novel coronavirus referred to as severe acute respiratory syndrome coronavirus 2 (SARS-CoV-2) was declared pandemic by the World Health Organization on 11^th^ March 2020.^1^ By 11^th^ May, in the absence of widespread testing in most countries, there was been global notification of 4 million confirmed cases of coronavirus disease 2019 (COVID-19) – the disease caused by SARS-CoV-2 – with it being implicated in more than 270,000 deaths.^2^ Equivalent data releases for the UK indicated 223,060 cases and 32,065 fatalities.^3^

Prior pandemics – Spanish influenza in 1918 and Swine influenza in 2009 – were notable for marked inequalities in their occurrence, whereby more socioeconomically disadvantaged countries,^4,5^ cities,^6^ neighbourhoods,^7,8^ and individuals^9^ experienced the highest mortality rates from the infection. Recent findings from analyses of data for COVID-19 hospitalisations across the five boroughs of New York City^10^ and deaths involving the infection in the UK^11^ reveal higher rates in more deprived areas. The mechanisms that underlie these differences are likely to be numerous and might involve overcrowded living and working conditions, comorbidity, poor access to healthcare, and a relative lack of understanding of prevention advice among socially disadvantaged individuals.^12^ Indirect pathways might include the higher prevalence of unfavourable health behaviours – cigarette smoking, alcohol intake, and suboptimal nutrition – in lower social groups which in themselves have been linked to selected lower respiratory tract infections.^13^

Whereas they are correlated with socioeconomic status,^14,15^ mental health and cognitive function might have independent utility in understanding the burden of respiratory disease. Poor mental health has been hypothesised to be a potential consequence of COVID-19 based on the findings of studies of survivors of the severe acute respiratory syndrome (SARS) pandemic,^16,17^ but it may also influence the risk of contracting the infection, at least in part by impairing innate or adaptive immunity ^18^ and diminishing the precautions taken to minimise risk. In a cross-sectional study, mental health problems were correlated with a higher likelihood of reporting the common cold,^19^ a coronavirus species. In cohort studies generated using linked electronic registries, people with a history of depression,^20^ psychosis,^21^ and stress disorders^22^ serious enough to warrant treatment in a psychiatric care facility subsequently experienced elevated rates of an array of respiratory infections. Additionally, in the general population, even moderate levels of self-reported symptoms of psychological distress (depression and anxiety) have been prospectively linked to an elevated risk of death from pneumonia despite adjustment for confounding factors which include socioeconomic position.^23^

In the COVID-19 pandemic, the public has been offered much preventative advice and guidelines which span the simple and practical to the complex, contradictory and false.^24-26^ In order to diminish their risk of the infection, the population has to acquire, synthesise, and deploy this information – described by some commentators as an ‘infodemic’ – but the ability to do so seems to vary by levels of health literacy^27^ just as it may for its close correlate, cognitive function. Although traditionally studied in the context of non-communicable disease,^28-30^ higher levels of cognitive ability – a psychological trait that involves the storage, selection, manipulation, and organisation of information – appear to be related to markedly lower rates of mortality from infectious disease after taking into account social circumstances.^31,32^

With this evidence base giving us reason to anticipate a relation of these socioeconomic and psychological characteristics with incident COVID-19 infection, we explored them using data from UK Biobank, a large prospective cohort study. To the best of our knowledge, this is the first examination of the role of individual-level psychosocial characteristics in the primary prevention of COVID-19.

## Methods

We used data from both UK Biobank, a prospective cohort study, the sampling and procedures of which have been well described.^33^ In brief, baseline data collection took place between 2006 and 2010 in twenty-two research assessment centres across the UK, resulting in a sample of 502,655 people aged 40 to 69 years (response rate 5.5%).^33^ In UK Biobank, ethical approval was received from the North-West Multicentre Research Ethics Committee, and the research was carried out in accordance with the Declaration of Helsinki of the World Medical Association, and participants gave informed consent. No additional ethical approval was required for present analyses of anonymised data.

### Assessment of socioeconomic factors

We used four indicators of socioeconomic status. Total annual household income before tax was self-reported and classified into three groups (<18,000, −30,999, −51,999, ≥£52,000 GBP). For educational qualifications, we used a three category variable (degree, other qualifications, no qualifications). Using Standard Occupational Classifications of current job, or most recent if participants were not working or data on current job were missing, we produced three categories with managerial positions having the highest prestige: managers & senior officials, professional, associate professional & technical; administrative & secretarial, & skilled; and personal service, sales & customer service, process, plant & machine operatives, elementary. Lastly, we used the Townsend deprivation index as our indicator of neighbourhood socioeconomic circumstances. Based on a composite of four characteristics (home and car ownership, employment, and number of household resident), participants’ postcodes at recruitment were matched to areas from the most recent national census. A continuously scored variable, higher values denote greater deprivation.

### Assessment of psychological factors

We used five psychological factors. Study members were asked if they had ever been under the care of a psychiatrist for any mental health problem; in the UK, such a referral would ordinarily have been triaged via a general practitioner. Symptoms of psychological distress – anxiety, worrying, anhedonia, and depression – were measured using the four item version of the Patient Health Questionnaire (PHQ-4)^6^ in which individual items are rated on a 4 point Likert scale from 0 (“not at all”) to 3 (“nearly every day”) such that total scores range from 0 to 12 (higher scores denote greater distress). Scores on the PHQ-4 show good agreement with longer scales, and reveal known correlations with demographic risk factors for depression and anxiety.^7^ Neuroticism was measured with the 12-item Eysenck Personality Questionnaire-Revised Short Form;^8^ higher scores denote higher levels.

Scores from two tests of cognitive functioning were used. Verbal and numerical reasoning was measured using a computerized 13-item multiple-choice test with a two-minute time limit. The score was the number of correct answers. This test was introduced after the beginning of the baseline assessment period so data are available for a subset of study members (N=180,914). Reaction time was measured using a computerized Go/No-Go “Snap” game. Participants were presented with electronic images of two cards. If symbols on the cards were identical, participants were instructed to immediately push the button-box using their dominant hand. The first five pairs were used as a practice with the remaining seven pairs, containing four identical cards, forming the assessment. Reaction time score was the mean time (milliseconds) to press the button when each of these four pairs was presented. Choice reaction time correlates strongly with single mental tests that involve complex reasoning and knowledge.^34^

### Assessment of confounding factors

Ethnicity was self-reported and categorised as White, Asian, Black, Chinese, Mixed, or other ethnic group. A social isolation scale was derived from enquiries concerning number of people in household, visiting friends/family, and social activities.^35^ One point was allocated for living alone, one for friends/family visits less than once/month, and one for no weekly participation in social activities. A dichotomous variable was derived with social isolation denoted by a score of 3. Self-reported physician diagnosis was collected for vascular or heart problems, diabetes, chronic lung disease, asthma, and cancer.

Cigarette smoking, physical activity, and alcohol consumption were measured using standard enquiries. Height and weight were measured directly during a medical examination from which body mass index was calculated using the usual formula (weight, kg/height,^2^ m^2^). Forced expiratory volume in one second, a measure of pulmonary function, was quantified using spirometry with the best of three technically satisfactory exhalations used in our analyses. Handgrip strength was measured using a hydraulic hand dynamometer (Jamar J00105) with the participant maximally squeezing the handle of the dynamometer while seated for 3 seconds; an average of the readings from the right and left hand was used. Seated systolic and diastolic blood pressure measurements were made twice using the Omron HEM-7015IT digital blood pressure monitor (Omron Healthcare)^20^ or, exceptionally, a manual sphygmomanometer; an average of the two readings was used herein. We defined hypertension according to existing guidelines as systolic/diastolic blood pressure ≥140/90 mmHg and/or self-reported use of antihypertensive medication.^36^ Non-fasting venous blood, available in a sub-sample, was drawn with assaying conducted at dedicated central laboratory for C-reactive protein, glycated haemoglobin A1c, and high-density lipoprotein cholesterol.^37^

### Ascertainment of hospitalisation for COVID-19

Provided by Public Health England, data on COVID-19 status downloaded on 1^st^ May 2020 covered the period 16^th^ March 2020 until 26^th^ April 2020.^38^ Nose and/or throat swabs were taken from hospitalised patients and detection of SARS-CoV-2 can be regarded as an indication of severe disease.^38^ With coverage being for England only, study members from Scotland and Wales were omitted from our analytical sample.

In preliminary analyses, we used three different COVID-19 case definitions based on these data: all apparent cases of the disease (N=908); cases based on samples from in-patients only (N=751); and cases based on two or more samples from in-patients (N=445) – the notion being that these patients were amongst the most severe cases. Evidence from prognostic studies of hospitalised patients in the USA^39^ and China^40^ suggest that men, older individuals, ethnic minorities, and those with existing disease experience greater rates of progression to intensive care and death. Preliminary analyses of the present data on incidence of severe disease revealed similar associations irrespective of case definition (supplemental table 1). On the basis of the similarity of this predictive validity, we proceeded with our main analyses in which we used all COVID-19 cases (N=908).

### Statistical analyses

We omitted from our analyses men and women who had died before 5^th^ March 2020 – the latest date to which vital status data were available – as they could not contribute to the risk set for COVID-19. Odds ratios and accompanying 95% confidence intervals were computed using logistic regression models to summarise the relationship between psychosocial factors and COVID-19 hospitalisations. In the main analyses, we initially adjusted odds ratios for age and sex, followed by ethnicity, then covariates organised into comorbidities (vascular disease, diabetes etc.), lifestyle factors (cigarette smoking etc.), and, depending on the psychosocial exposures of interest, socioeconomic or psychological factors. In preliminary analyses, the addition of biomarkers to the final model had no appreciable impact on the effects estimates relative to the final model in which they did not feature (supplemental tables 4 and 5 versus tables 2 and 3); these covariates were therefore not included in the main analyses. Analyses were conducted using Stata version 13.

### Patient involvement

These analyses are based on existing data of a typically healthy population. We were not involved in their recruitment; thus, to our knowledge, no patients were explicitly engaged in designing the present research question or the outcome measures, nor were they involved in developing plans for recruitment, design, or implementation of the study. No patients were asked to advise on interpretation or writing up of results. Results from UK Biobank are routinely disseminated to study participants via the study website and social media outlets.

## Results

In 431,051 study members (236,725 women) there were 908 hospitalisations for COVID-19 between 16 March 2020 and 26^th^ April in England (402 in women). Of the 28 baseline characteristics featured in table 1, only four – extant cancer, grip strength, neuroticism, and social isolation – did not reveal relationships with COVID-19 at conventional levels of statistical significance in unadjusted analyses. These were therefore excluded as covariates from subsequent multiple regression analyses.

In table 2 and figure 1 we depict the association between various socio-economic characteristics and the risk of hospitalisation for COVID-19 infection. After adjustment for age and sex, those study members who were most disadvantaged educationally, financially, and geographically experienced around a doubling in the risk of infection. Effects in these analyses were apparent across the full socioeconomic continuum (p for trend <0.0001). Whereas controlling for ethnicity had little impact on these gradients, partial attenuation was apparent after taking into account comorbidities and lifestyle factors.

**Table 1.**
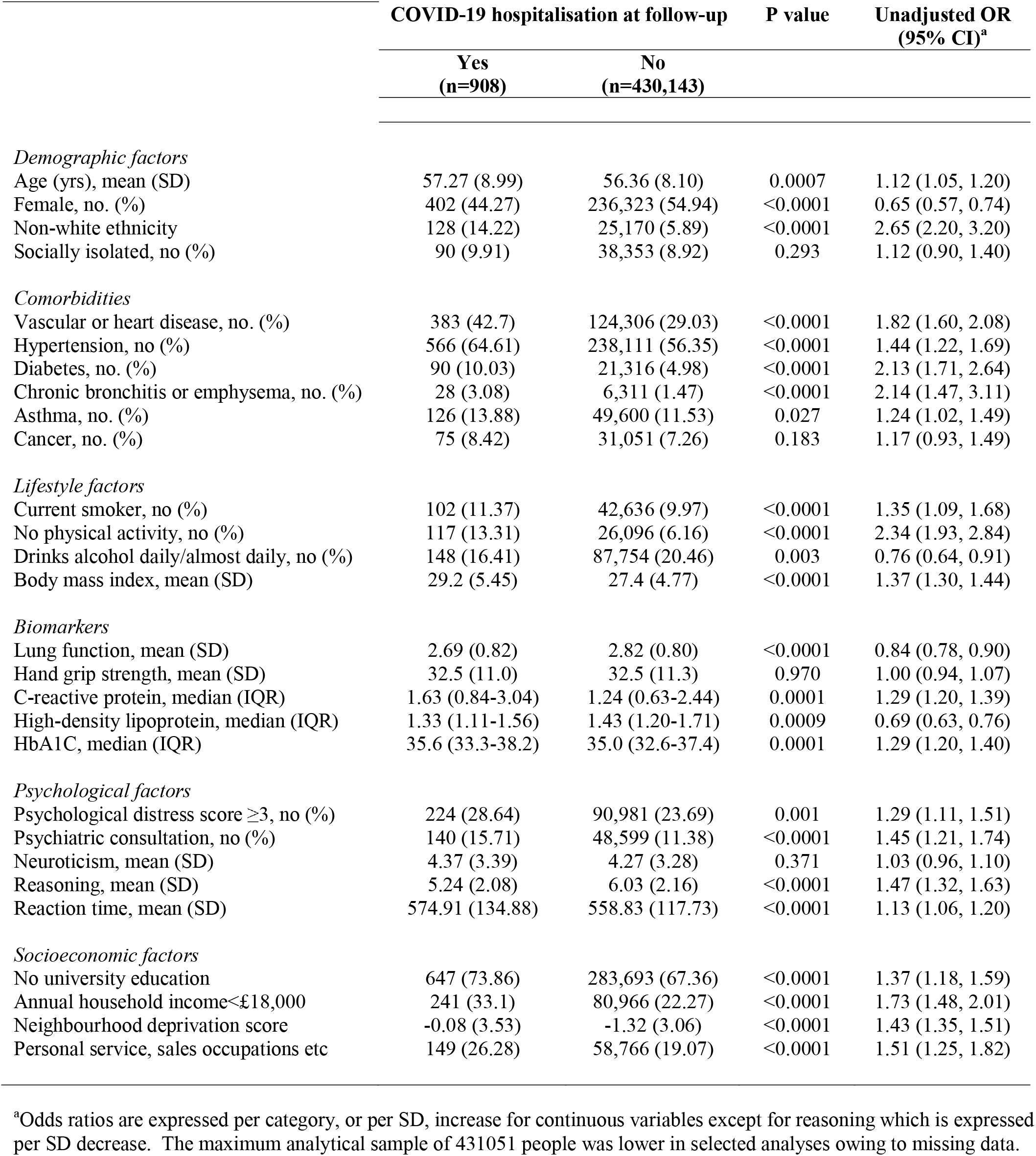
Psychosocial factors and covariates at baseline according to hospitalisations for COVID-19

|  | COVID-19 hospitalisation at follow-up |  | P value | Unadjusted OR<br>(95% CI)<sup>a</sup> |
| --- | --- | --- | --- | --- |
|  | Yes<br>(n=908) | No<br>(n=430,143) |  |  |
| *Demographic factors* |  |  |  |  |
| Age (yrs), mean (SD) | 57.27 (8.99) | 56.36 (8.10) | 0.0007 | 1.12 (1.05, 1.20) |
| Female, no. (%) | 402 (44.27) | 236,323 (54.94) | <0.0001 | 0.65 (0.57, 0.74) |
| Non-white ethnicity | 128 (14.22) | 25,170 (5.89) | <0.0001 | 2.65 (2.20, 3.20) |
| Socially isolated, no (%) | 90 (9.91) | 38,353 (8.92) | 0.293 | 1.12 (0.90, 1.40) |
| *Comorbidities* |  |  |  |  |
| Vascular or heart disease, no. (%) | 383 (42.7) | 124,306 (29.03) | <0.0001 | 1.82 (1.60, 2.08) |
| Hypertension, no (%) | 566 (64.61) | 238,111 (56.35) | <0.0001 | 1.44 (1.22, 1.69) |
| Diabetes, no. (%) | 90 (10.03) | 21,316 (4.98) | <0.0001 | 2.13 (1.71, 2.64) |
| Chronic bronchitis or emphysema, no. (%) | 28 (3.08) | 6,311 (1.47) | <0.0001 | 2.14 (1.47, 3.11) |
| Asthma, no. (%) | 126 (13.88) | 49,600 (11.53) | 0.027 | 1.24 (1.02, 1.49) |
| Cancer, no. (%) | 75 (8.42) | 31,051 (7.26) | 0.183 | 1.17 (0.93, 1.49) |
| *Lifestyle factors* |  |  |  |  |
| Current smoker, no (%) | 102 (11.37) | 42,636 (9.97) | <0.0001 | 1.35 (1.09, 1.68) |
| No physical activity, no (%) | 117 (13.31) | 26,096 (6.16) | <0.0001 | 2.34 (1.93, 2.84) |
| Drinks alcohol daily/almost daily, no (%) | 148 (16.41) | 87,754 (20.46) | 0.003 | 0.76 (0.64, 0.91) |
| Body mass index, mean (SD) | 29.2 (5.45) | 27.4 (4.77) | <0.0001 | 1.37 (1.30, 1.44) |
| *Biomarkers* |  |  |  |  |
| Lung function, mean (SD) | 2.69 (0.82) | 2.82 (0.80) | <0.0001 | 0.84 (0.78, 0.90) |
| Hand grip strength, mean (SD) | 32.5 (11.0) | 32.5 (11.3) | 0.970 | 1.00 (0.94, 1.07) |
| C-reactive protein, median (IQR) | 1.63 (0.84-3.04) | 1.24 (0.63-2.44) | 0.0001 | 1.29 (1.20, 1.39) |
| High-density lipoprotein, median (IQR) | 1.33 (1.11-1.56) | 1.43 (1.20-1.71) | 0.0009 | 0.69 (0.63, 0.76) |
| HbA1C, median (IQR) | 35.6 (33.3-38.2) | 35.0 (32.6-37.4) | 0.0001 | 1.29 (1.20, 1.40) |
| *Psychological factors* |  |  |  |  |
| Psychological distress score ≥3, no (%) | 224 (28.64) | 90,981 (23.69) | 0.001 | 1.29 (1.11, 1.51) |
| Psychiatric consultation, no (%) | 140 (15.71) | 48,599 (11.38) | <0.0001 | 1.45 (1.21, 1.74) |
| Neuroticism, mean (SD) | 4.37 (3.39) | 4.27 (3.28) | 0.371 | 1.03 (0.96, 1.10) |
| Reasoning, mean (SD) | 5.24 (2.08) | 6.03 (2.16) | <0.0001 | 1.47 (1.32, 1.63) |
| Reaction time, mean (SD) | 574.91 (134.88) | 558.83 (117.73) | <0.0001 | 1.13 (1.06, 1.20) |
| *Socioeconomic factors* |  |  |  |  |
| No university education | 647 (73.86) | 283,693 (67.36) | <0.0001 | 1.37 (1.18, 1.59) |
| Annual household income<£18,000 | 241 (33.1) | 80,966 (22.27) | <0.0001 | 1.73 (1.48, 2.01) |
| Neighbourhood deprivation score | -0.08 (3.53) | -1.32 (3.06) | <0.0001 | 1.43 (1.35, 1.51) |
| Personal service, sales occupations etc | 149 (26.28) | 58,766 (19.07) | <0.0001 | 1.51 (1.25, 1.82) |

<sup>a</sup>Odds ratios are expressed per category, or per SD, increase for continuous variables except for reasoning which is expressed per SD decrease. The maximum analytical sample of 431051 people was lower in selected analyses owing to missing data.

**Table 2.** Odds ratios (95% CI) for the relation of socioeconomic factors with COVID-19 hospitalisation

|  | Case no./Risk no.<sup>1</sup> | Adjustments |  |  |  |  |  |
| --- | --- | --- | --- | --- | --- | --- | --- |
|  |  | Age & sex | Age, sex & ethnicity | Age, sex, ethnicity & comorbidity<sup>2</sup> | Age, sex, ethnicity & lifestyle factors<sup>3</sup> | Age, sex, ethnicity & psychological factors<sup>4</sup> | All covariates |
| **Educational attainment** |  | N=422057 | N=420502 | N=415945 | N=415367 | N=155244 | N=152739 |
| University degree | 229/137717 | 1.0 (ref) | 1.0 (ref) | 1.0 (ref) | 1.0 (ref) | 1.0 (ref) | 1.0 (ref) |
| Other qualifications | 406/214337 | 1.16 (0.98, 1.36) | 1.19 (1.01, 1.41) | 1.16 (0.98, 1.37) | 1.03 (0.87, 1.22) | 1.06 (0.81, 1.39) | 0.94 (0.71, 1.24) |
| No qualifications | 241/70003 | 2.05 (1.70, 2.47) | 2.07 (1.71, 2.50) | 1.85 (1.53, 2.25) | 1.47 (1.20, 1.80) | 1.35 (0.93, 1.95) | 1.01 (0.68, 1.49) |
| P for trend |  | <0.0001 | <0.0001 | <0.0001 | <0.0001 | 0.151 | 0.945 |
| **Annual household income** |  | N=364219 | N=363175 | N=359853 | N=359491 | N=137808 | N=135773 |
| <£18,000 | 241/81207 | 2.00 (1.63, 2.47) | 1.89 (1.51, 2.35) | 1.74 (1.39, 2.17) | 1.39 (1.10, 1.75) | 1.34 (0.91, 1.97) | 1.15 (0.77, 1.73) |
| £18,000-£30,999 | 179/92461 | 1.31 (1.05, 1.63) | 1.27 (1.01, 1.60) | 1.22 (0.97, 1.54) | 1.05 (0.83, 1.32) | 1.29 (0.90, 1.85) | 1.15 (0.79, 1.68) |
| £31,000-£51,999 | 167/95454 | 1.18 (0.94, 1.48) | 1.17 (0.94, 1.47) | 1.16 (0.92, 1.45) | 1.07 (0.85, 1.34) | 1.03 (0.72, 1.49) | 1.02 (0.70, 1.48) |
| ≥£52,000 | 141/95097 | 1.0 (ref) | 1.0 (ref) | 1.0 (ref) | 1.0 (ref) | 1.0 (ref) | 1.0 (ref) |
| P for trend |  | <0.0001 | <0.0001 | <0.0001 | 0.006 | 0.077 | 0.401 |
| **Neighbourhood deprivation** |  | N=430538 | N=427986 | N=419593 | N=418942 | N=156360 | N=153384 |
| 1 (low) | 205/143483 | 1.0 (ref) | 1.0 (ref) | 1.0 (ref) | 1.0 (ref) | 1.0 (ref) | 1.0 (ref) |
| 2 | 267/143548 | 1.32 (1.10, 1.58) | 1.29 (1.07, 1.55) | 1.25 (1.04, 1.50) | 1.20 (1.00, 1.45) | 1.32 (0.97, 1.79) | 1.22 (0.89, 1.65) |
| 3 | 436/143517 | 2.20 (1.86, 2.59) | 1.97 (1.66, 2.34) | 1.79 (1.51, 2.13) | 1.57 (1.31, 1.88) | 1.52 (1.12, 2.05) | 1.20 (0.87, 1.63) |
| P for trend |  | <0.0001 | <0.0001 | <0.0001 | <0.0001 | 0.007 | 0.297 |
| **Occupational classification** |  | N=308689 | N=307262 | N=302239 | N=302495 | N=130238 | N=128079 |
| Managers, senior officials, etc | 324/175637 | 1.0 (ref) | 1.0 (ref) | 1.0 (ref) | 1.0 (ref) | 1.0 (ref) | 1.0 (ref) |
| Administrative, secretarial, etc | 94/74137 | 0.70 (0.55, 0.88) | 0.69 (0.55, 0.87) | 0.68 (0.54, 0.86) | 0.63 (0.50, 0.80) | 0.65 (0.46, 0.92) | 0.59 (0.42, 0.84) |
| Personal service, sales, etc | 149/58915 | 1.39 (1.14, 1.69) | 1.30 (1.07, 1.59) | 1.22 (1.00, 1.49) | 1.06 (0.86, 1.30) | 0.90 (0.65, 1.26) | 0.75 (0.53, 1.06) |
| P for trend |  | 0.024 | 0.091 | 0.314 | 0.780 | 0.242 | 0.027 |

<sup>1</sup> Numbers based on unadjusted model. <sup>2</sup> Comorbidity includes diagnoses of vascular or heart disease, diabetes, chronic bronchitis or emphysema, asthma, and hypertension defined according to measured blood pressure and/or use of anti-hypertensive medication. <sup>3</sup> Lifestyle factors includes body mass index, smoking status, alcohol intake frequency & number of types of physical activity taken in last four weeks. <sup>4</sup> Psychological factors include psychological distress, psychiatric consultation, neuroticism, verbal and numerical reasoning, & reaction time.

**Table 3.** Odds ratios (95% CI) for the relation of psychological factors with COVID-19 hospitalisation

|  | Case no./<br>Risk no.<sup>1</sup> | Adjustments |  |  |  |  |  |
| --- | --- | --- | --- | --- | --- | --- | --- |
|  |  | Age & sex | Age, sex &<br>ethnicity | Age, sex,<br>ethnicity &<br>comorbidity<sup>2</sup> | Age, sex,<br>ethnicity &<br>lifestyle factors<sup>3</sup> | Age, sex,<br>ethnicity &<br>socioeconomic<br>factors<sup>4</sup> | All covariates |
| **Psychological distress** |  | N=384909 | N=383655 | N=377290 | N=376562 | N=248162 | N=245119 |
| 1 (low) | 267/153504 | 1.0 (ref) | 1.0 (ref) | 1.0 (ref) | 1.0 (ref) | 1.0 (ref) | 1.0 (ref) |
| 2 | 291/140200 | 1.28 (1.08, 1.51) | 1.29 (1.09, 1.53) | 1.22 (1.03, 1.45) | 1.16 (0.98, 1.38) | 1.16 (0.93, 1.45) | 1.07 (0.86, 1.35) |
| 3 | 224/91205 | 1.58 (1.32, 1.89) | 1.51 (1.26, 1.81) | 1.37 (1.14, 1.65) | 1.18 (0.98, 1.43) | 1.26 (0.98, 1.61) | 1.09 (0.84, 1.41) |
| P for trend |  | <0.0001 | <0.0001 | 0.001 | 0.068 | 0.064 | 0.487 |
| Per SD increase |  | 1.22 (1.14, 1.29) | 1.19 (1.12, 1.26) | 1.15 (1.08, 1.23) | 1.09 (1.02, 1.17) | 1.12 (1.02, 1.22) | 1.07 (0.97, 1.17) |
| **Psychiatric consultation** |  | N=427819 | N=426823 | N=418218 | N=417481 | N=269373 | N=265566 |
| No | 751/379080 | 1.0 (ref) | 1.0 (ref) | 1.0 (ref) | 1.0 (ref) | 1.0 (ref) | 1.0 (ref) |
| Yes | 140/487739 | 1.50 (1.25, 1.79) | 1.51 (1.26, 1.81) | 1.45 (1.21, 1.75) | 1.32 (1.09, 1.59) | 1.23 (0.94, 1.62) | 1.15 (0.87, 1.52) |
| **Neuroticism** |  | N=425707 | N=424212 | N=416378 | N=415622 | N=265538 | N=264784 |
| 1 (low) | 224/106910 | 1.0 (ref) | 1.0 (ref) | 1.0 (ref) | 1.0 (ref) | 1.0 (ref) | 1.0 (ref) |
| 2 | 345/174705 | 1.01 (0.85, 1.19) | 1.03 (0.87, 1.22) | 0.99 (0.83, 1.17) | 0.99 (0.83, 1.18) | 1.03 (0.82, 1.30) | 1.01 (0.80, 1.27) |
| 3 | 319/144092 | 1.19 (1.00, 1.42) | 1.21 (1.02, 1.44) | 1.10 (0.92, 1.31) | 1.08 (0.90, 1.29) | 1.06 (0.84, 1.35) | 1.00 (0.78, 1.28) |
| P for trend |  | 0.038 | 0.023 | 0.277 | 0.382 | 0.621 | 0.985 |
| Per SD increase |  | 1.08 (1.01, 1.15) | 1.08 (1.01, 1.16) | 1.05 (0.98, 1.12) | 1.03 (0.96, 1.10) | 1.00 (0.92, 1.10) | 0.99 (0.90, 1.09) |
| **Verbal numerical reasoning** |  | N=175267 | N=174581 | N=172530 | N=415777 | N=126721 | N=124890 |
| 1 (low) | 152/43988 | 2.66 (2.06, 3.34) | 2.31 (1.77, 3.02) | 2.17 (1.65, 2.86) | 1.92 (1.45, 2.53) | 2.14 (1.50, 3.05) | 1.98 (1.38, 2.85) |
| 2 | 115/58446 | 1.52 (1.16, 1.99) | 1.45 (1.10, 1.90) | 1.46 (1.10, 1.92) | 1.36 (1.03, 1.80) | 1.57 (1.14, 2.17) | 1.58 (1.14, 2.18) |
| 3 | 96/72833 | 1.0 (ref) | 1.0 (ref) | 1.0 (ref) | 1.0 (ref) | 1.0 (ref) | 1.0 (ref) |
| P for trend |  | <0.0001 | <0.0001 | <0.0001 | <0.0001 | <0.0001 | <0.0001 |
| Per SD decrease |  | 1.47 (1.32, 1.64) | 1.37 (1.23, 1.53) | 1.33 (1.19, 1.49) | 1.27 (1.13, 1.42) | 1.35 (1.17, 1.57) | 1.31 (1.13, 1.52) |
| **Reaction time** |  | N=426147 | N=424432 | N=416366 | N=415777 | N=268826 | N=265002 |
| 1 (low) | 262/140934 | 1.0 (ref) | 1.0 (ref) | 1.0 (ref) | 1.0 (ref) | 1.0 (ref) | 1.0 (ref) |
| 2 | 274/141575 | 1.04 (0.87, 1.23) | 1.00 (0.84, 1.19) | 0.97 (0.82, 1.16) | 0.95 (0.79, 1.13) | 1.02 (0.82, 1.27) | 1.02 (0.81, 1.27) |
| 3 | 345/143368 | 1.27 (1.08, 1.51) | 1.16 (0.98, 1.37) | 1.11 (0.93, 1.32) | 1.04 (0.88, 1.24) | 1.06 (0.84, 1.34) | 1.02 (0.80, 1.29) |
| P for trend |  | 0.004 | 0.078 | 0.205 | 0.572 | 0.608 | 0.876 |
| Per SD increase |  | 1.12 (1.06, 1.19) | 1.07(1.01, 1.14) | 1.06 (0.99, 1.13) | 1.03 (0.97, 1.10) | 1.08 (0.98, 1.18) | 1.07 (0.97, 1.17) |

<sup>1</sup> Numbers based on age & sex adjusted model. <sup>2</sup> Comorbidity includes diagnoses of vascular or heart disease, diabetes, chronic bronchitis or emphysema, asthma, and hypertension defined according to measured blood pressure and/or use of anti-hypertensive drugs. <sup>3</sup> Lifestyle factors included body mass index, smoking status, alcohol intake frequency & number of types of physical activity taken in last four weeks.
<sup>4</sup> Socioeconomic factors included occupational classification, educational attainment, Townsend deprivation index, & household income before tax

**Figure 1.**
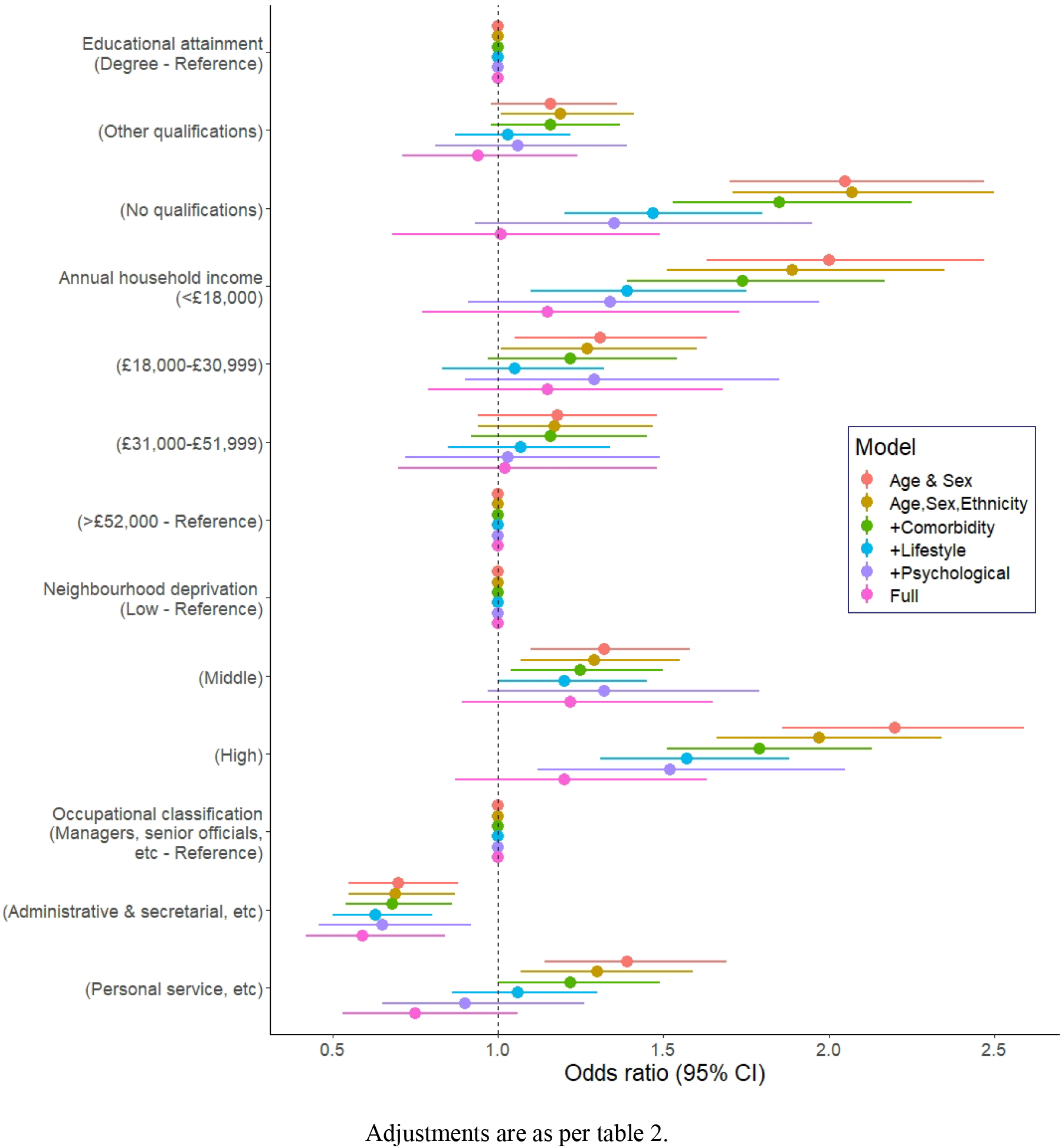
Odds ratios (95% CI) for the relation of socioeconomic factors with COVID-19 hospitalisation

However, adjusting for psychological characteristics had the largest attenuating effect relative to the minimally-adjusted (age, sex, and ethnicity) odds ratios. Although the risk of hospitalisation remained somewhat elevated at both lower levels of education and income, statistical significance at conventional levels was lost. Given the known correlation between education and cognitive ability (herein, r=-0.40, p-value <0.0001), in sensitivity analyses we removed verbal and numerical reasoning test scores from the model containing the 5 psychological factors. This resulted in the magnitude of the low education– COVID-19 relationship being restored (odds ratio; 95% confidence interval for no qualifications: 2.08; 1.69, 2.56) and suggested most of the marked attention seen for this relationship after taking into account psychological factors could be ascribed to individual differences in cognitive ability rather than education. The association between area deprivation and risk of infection was more robust to these various statistical adjustments.

Of the socioeconomic variables, occupational classification of the study members revealed the weakest association with hospitalisation for COVID-19 and, in all analyses, study members in the administrative/secretarial occupations in fact experienced some protection against the infection. Lastly, after including up to seventeen covariates in the most complex multivariable models, there was evidence of some weak residual associations for income and deprivation but not for education.

In table 2 and figure 2 we illustrate the associations between psychological traits and the risk of COVID-19. In minimally-adjusted (age, sex, and ethnicity) analyses, all five psychological factors were related to the risk of hospitalisation with the infection. Effects for neuroticism and reaction time – weak initially – were essentially eliminated after control for comorbidities and any subsequent group of covariates. Adjustment for comorbidities also had a partial impact on the relation of distress, psychiatric consultation, and verbal and numerical reasoning with the infection, but associations largely remained, most obviously for reasoning score. After multiple control for all covariates, however, the only relationship that remained with COVID-19 was that for verbal and numerical reasoning such that the most disadvantaged group experienced around a doubling of hospitalisation risk.

**Figure 2.**
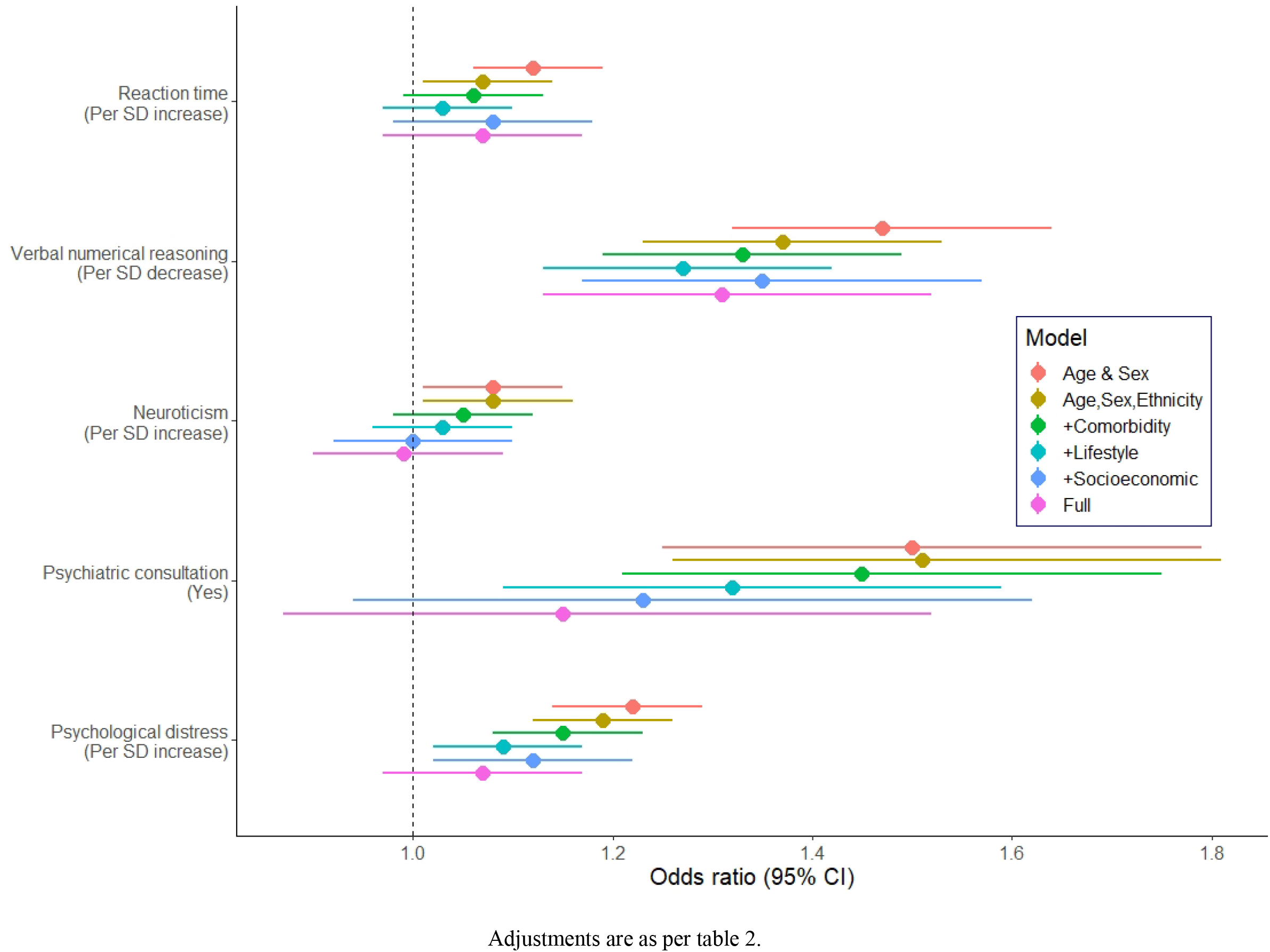
Odds ratios (95% CI) for the relation of psychological factors with COVID-19 hospitalisation

We also carried out some planned sensitivity analyses. With the verbal and numerical reasoning test having been introduced part way through baseline data collection, as indicated, analyses featuring this variable were based on a subgroup of study members. To ensure direct comparison across statistical models, for each exposure we therefore recomputed our analyses based on a non-missing dataset (supplemental table 2 for socioeconomic characteristics, and supplemental table 3 for psychological characteristics). The same patterns of association was apparent in these sensitivity analyses.

## Discussion

### Principal findings

Our main findings were that disadvantaged levels of a series of psychosocial characteristics – education, income, area deprivation, mental health, and cognitive function – were related to an elevated risk of hospitalisations with COVID-19 in most of the analyses conducted. Net of mutual control for these factors, and after taking into account several other potential confounders, however, only the association of lower cognitive function based on a test of verbal and numerical reasoning with a higher risk of this infection remained. That we were able to replicate findings for apparently known risk factors for COVID-19 from prognostic studies – being male, having an ethnic minority background, carrying a comorbidity – provides some support for the more novel findings for these psychosocial factors.

Our finding that the intermediate occupational group experienced a lower risk of hospitalisation was unexpected. A *post hoc* explanation is that this apparent ‘J’-shaped relation could in part be driven by the higher prestige category containing some medical professionals and, at the opposite end of the continuum, the personal services group being partially composed of carers, both of whom would be more likely to be exposed to the virus. Cardiovascular disease death is an exemplar of socioeconomic inequalities in disease risk,^41^ and analyses of this endpoint in relation to these occupational groups revealed a graded effect such that a doubling of risk was apparent in the most disadvantage group (age- and sex-adjusted hazard ratio; 95% confidence interval: 1.90; 1.61, 2.25) with intermediate rates evident in the administrative class (1.32; 1.11, 1.58). As such, these occupational classifications have some predictive validity, so lending some support to the apparently surprising result for COVID-19.

### Comparison with results from other studies

We are unaware of any published studies exploring the impact of individual-level psychosocial factors on the occurrence of risk of COVID-19. Prognostic studies using area-based statistics have recently been published, however. In New York City, Manhattan, the most socioeconomically advantaged borough based on routinely collected education and poverty statistics, had the lowest rates of hospitalisations for COVID-19 relative to the four remaining areas.^10^ While, by contrast, the Bronx, the least favourable socioeconomically, had the highest level of hospitalisations, rates were graded across the boroughs for education but not poverty. In a recent report from the Office for National statistics in the UK, rates of death in which COVID-19 was implicated were directly related to neighbourhood deprivation in a stepwise manner.^11^ Outside the eras of pandemics, other respiratory diseases such as tuberculosis,^42,43^ pneumonia,^44^ influenza,^45^ and, importantly, the common cold^46^ – also appear to be similarly socioeconomically patterned, although these are not universal observations.^47,48^ We are unaware of any studies exploring the relation of indicators of cognitive function and mental health with COVID-19, though up to a doubling in rates of death from respiratory disease has been reported in people with lower cognition test scores,^31,32^ individuals with a serious mental illness,^21^ and those with higher levels of psychological distress.^23^

### Mechanisms of effect

Specific and non-specific mechanisms may link these psychosocial variables to the risk of COVID-19. A plausible explanation for the association between cognition and respiratory infection is that people with higher ability, and indeed the educationally advantaged,^12^ may be more likely to take-up influenza and pneumococcal inoculation; however, in absence of any effective vaccination for COVID-19 this is implausible. In our analyses we took into account unfavourable health behaviours which are more common in lower cognition scoring groups^49-53^ and have also been implicated in the occurrence of pneumonia,^54^ but the effect for cognition remained. It may be that the deluge of health advice in the current pandemic during a period when news outlets and social media platforms have never been more ubiquitous, has highlighted that lower cognition and therefore poor health literacy in the population is a public health concern.^55,56^ In a small-scale cross-sectional study, people with low health literacy also reported being less concerned about the current pandemic and to believe they were at lower risk.^27^

Mental health problems may influence the risk of acquiring a respiratory infection by negatively impacting cognitive function,^57^ potentially compromising the ability to effectively take precautions to minimise the risk, adequately recognise a deterioration in health, actively seek medical attention, and communicate effectively with health care professionals. An unhealthy lifestyle and sub-optimal circumstances including poor housing and lower income are also more common in people with mental health problems^14,58^ but we were careful to covary on these factors in our analyses. It could also be the case that people experiencing higher levels of psychological distress have diminished learned resistance to infection owing to fewer social interactions, although a pre-pandemic measure of social isolation in our analyses did not confer the expected protection against the infection.

More speculative explanations for some of the effects found herein are that our outcome, hospitalisation for COVID-19 infection, represents not only the occurrence of the infection itself but also a sub-optimal viral-response. People with a higher burden of psychological distress – which includes worry about physical health – might be more concerned about becoming seriously unwell and therefore have a lower symptom severity threshold for visiting hospital. Similarly, individuals with lower cognition may have less confidence in their own decision-making, self-care, and UK government messages to remain at home when unwell, instead resorting to hospital-based advice.

### Study strengths and limitations

The strengths of our study include it being well characterised for exposures and covariates despite its scale, allowing us to attempt to identify independent effects. That the study is prospective means assessment of these baseline data preceded that of disease onset; as such, reverse causality is not a concern such that the infection could not, for instance, influence mental health and job loss leading to downward social mobility. Our work has its weaknesses. Samples were taken from hospitalised patients but it is unclear if all cases had been exclusively hospitalised because of COVID-19-type symptoms, or, as seem likely given mass testing within hospitals, some patients were found to be positive for the infection while an inpatient for other reasons. Our outcome also represents an unfavourable response to a viral challenge as opposed to disease incidence across the full population; the latter could only be ascertained with comprehensive testing of our study sample or indeed the population of England as a whole. We excluded study members who had died prior to 5^th^ March 2020 because they could not contribute to the risk set, however, ascertainment of hospitalisations for COVID-19 did not reliably begin until 16^th^ March. It is unlikely, however, that the absence of vital status data for this 11 day period would have substantially biased our effect estimates in this large dataset.

The UK Biobank study sample is recruited from only 5.5% of the target population agreeing to participate.^33^ As has been demonstrated,^59,60^ the data material is therefore inappropriate for estimation of risk factor or disease prevalence and incidence of COVID-19 infection, and any data simulations of its dissemination. These observations do not, however, seem to influence reproducibility of the association of established risk factors for non-communicable disease such as vascular disease and selected cancers, and other health endpoints such as suicide.^60^ We think the same reasoning can be applied to associations with communicable diseases.

### Conclusions

In conclusion, in aetiological-orientated analyses of data from this prospective cohort study, a range of psychosocial factors showed associations with subsequent hospitalisations for COVID-19, among which cognitive function – a potential marker of health literacy – was most robustly related. These findings have important implications for public health messaging, but replication is required before policy recommendations can be advanced.

## Data Availability

Data from UK Biobank (http://www.ukbiobank.ac.uk/) are available to bona fide researchers upon application.

## Acknowledgement

We thank UK Biobank study members for their generosity in participating.

## Access to data

Data from UK Biobank (http://www.ukbiobank.ac.uk/) are available to *bona fide* researchers upon application. Part of this research has been conducted using the UK Biobank Resource under Application 10279.

## Transparency

GDB affirms that the manuscript is an honest, accurate, and transparent account of the study being reported; that no important aspects of the study have been omitted; and that any discrepancies from the study as planned have been explained.

## Contributions

The authors collectively generated the idea for the present paper and formulated an analytical plan; ML built the dataset; CRG carried out the data analyses; DMA prepared the figures; and GDB wrote the first draft of manuscript. All authors commented on an earlier version of the manuscript. GDB, CRG, ML and IJD will act as guarantors for this work. The corresponding author attests that all listed authors meet authorship criteria and that no others meeting the criteria have been omitted. CRG and ML had full access to UK Biobank data. GDB takes responsibility for the decision to submit the manuscript for publication.

## Dissemination to participants and related patient and public communities

Findings will be disseminated via the media departments of the authors’ institutes. Results from UK Biobank are routinely disseminated to study participants via the study website and Twitter feed.

## Ethical approval

In UK Biobank, ethical approval for data collection was received from the North-West Multi-centre Research Ethics Committee and the research was carried out in accordance with the Declaration of Helsinki of the World Medical Association.

## Open access statement

University College London will cover open access charges (subject to confirmation).

## Exclusive licence

The Corresponding Author has the right to grant on behalf of all authors and does grant on behalf of all authors, a worldwide licence (http://www.bmj.com/sites/default/files/BMJ%20Author%20Licence%20March%202013.doc) to the Publishers and its licensees in perpetuity, in all forms, formats and media (whether known now or created in the future), to i) publish, reproduce, distribute, display and store the Contribution, ii) translate the Contribution into other languages, create adaptations, reprints, include within collections and create summaries, extracts and/or, abstracts of the Contribution and convert or allow conversion into any format including without limitation audio, iii) create any other derivative work(s) based in whole or part on the on the Contribution, iv) to exploit all subsidiary rights to exploit all subsidiary rights that currently exist or as may exist in the future in the Contribution, v) the inclusion of electronic links from the Contribution to third party material where-ever it may be located; and, vi) licence any third party to do any or all of the above. All research articles will be made available on an open access basis (with authors being asked to pay an open access fee—see http://www.bmj.com/about-bmj/resources-authors/forms-policies-and-checklists/copyright-open-access-and-permission-reuse). The terms of such open access shall be governed by a Creative Commons licence—details as to which Creative Commons licence will apply to the research article are set out in our worldwide licence referred to above.

---

**Supplemental Table 1.** Analyses of apparent known risk factors (based on clinical studies) for COVID-19 hospitalisation in UK Biobank (N=431,052)

|  | **All COVID-19 cases**<br>**(N=908)** | **COVID-19 cases**<br>**known to have been**<br>**hospitalised**<br>**(‘origin’=1) (N=751)** | **COVID-19 cases**<br>**known to have been**<br>**hospitalised**<br>**(‘origin’=1) \*and\***<br>**with >=2 tests**<br>**conducted (N=445)** |
| --- | --- | --- | --- |
| *Age* |  |  |  |
| 40-49 | Ref (1.0) | Ref (1.0) | Ref (1.0) |
| 50-59 | 0.69 (0.57, 0.83) | 0.77 (0.63, 0.95) | 0.81 (0.62, 1.05) |
| 60+ | 1.09 (0.93, 1.27) | 1.16 (0.97, 1.38) | 1.17 (0.83, 1.48) |
| Per decade increase | 1.15 (1.06, 1.25) | 1.18 (1.08, 1.29) | 1.17 (1.04, 1.32) |
| *Sex* |  |  |  |
| Female | Ref (1.0) | Ref (1.0) | Ref (1.0) |
| Male | 1.53 (1.35, 1.75) | 1.56 (1.35, 1.81) | 1.55 (1.28, 1.87) |
| *Ethnicity* |  |  |  |
| White | Ref (1.0) | Ref (1.0) | Ref (1.0) |
| Non-white | 2.65 (2.20, 3.20) | 2.45 (1.98, 3.03) | 3.12 (2.42, 4.01) |
| *Long-standing illness* |  |  |  |
| No | Ref (1.0) |  |  |
| Yes | 2.06 (1.80, 2.35) | 2.03 (1.75, 2.04) | 2.26 (1.87, 2.73) |

**Supplemental Table 2.** Odds ratios (95% CI) for the relation of socioeconomic factors with COVID-19 hospitalisation *– based on complete data*

|  | Case no./<br>Risk no.<sup>1</sup> | Adjustments |  |  |  |  |  |
| --- | --- | --- | --- | --- | --- | --- | --- |
|  |  | Age & sex | Age, sex & ethnicity | Age, sex, ethnicity & comorbidity<sup>1</sup> | Age, sex, ethnicity & lifestyle factors<sup>2</sup> | Age, sex, ethnicity & psychological factors<sup>3</sup> | Adjusted for all covariates |
| **Educational attainment** |  |  |  |  |  |  |  |
| University Degree | 90/55905 | 1.0 (ref) | 1.0 (ref) | 1.0 (ref) | 1.0 (ref) | 1.0 (ref) | 1.0 (ref) |
| Other qualifications | 144/77597 | 1.17 (0.89, 1.52) | 1.19 (0.91, 1.55) | 1.15 (0.88, 1.49) | 1.03 (0.79, 1.34) | 1.06 (0.81, 1.39) | 0.94 (0.71, 1.24) |
| No qualifications | 54/19237 | 1.70 (1.20, 2.41) | 1.75 (1.23, 2.48) | 1.60 (1.12, 2.27) | 1.28 (0.89, 1.84) | 1.30 (0.89, 1.90) | 1.01 (0.68, 1.49) |
| P for trend |  | 0.005 | 0.003 | 0.015 | 0.245 | 0.214 | 0.945 |
| **Annual household income** |  |  |  |  |  |  |  |
| <£18,000 | 68/26578 | 1.90 (1.31, 2.77) | 1.79 (1.23, 2.60) | 1.63 (1.12, 2.38) | 1.41 (0.96, 2.08) | 1.41 (0.95, 2.09) | 1.15 (0.77, 1.73) |
| £18,000-£30,999 | 71/34301 | 1.52 (1.06, 2.20) | 1.47 (1.02, 2.11) | 1.41 (0.98, 2.03) | 1.28 (0.89, 1.85) | 1.28 (0.88, 1.86) | 1.15 (0.79, 1.68) |
| £31,000-£51,999 | 60/36526 | 1.19 (0.82, 1.72) | 1.16 (0.80, 1.68) | 1.14 (0.79, 1.65) | 1.07 (0.74, 1.56) | 1.09 (0.75, 1.57) | 1.02 (0.70, 1.48) |
| ≥£52,000 | 54/38368 | 1.0 (ref) | 1.0 (ref) | 1.0 (ref) | 1.0 (ref) | 1.0 (ref) | 1.0 (ref) |
| P for trend |  | <0.0001 | 0.001 | 0.006 | 0.053 | 0.060 | 0.401 |
| **Townsend Deprivation Index** |  |  |  |  |  |  |  |
| 1 (low) | 69/48529 | 1.0 (ref) | 1.0 (ref) | 1.0 (ref) | 1.0 (ref) | 1.0 (ref) | 1.0 (ref) |
| 2 | 102/53718 | 1.34 (0.99, 1.82) | 1.31 (0.97, 1.78) | 1.29 (0.95, 1.75) | 1.24 (0.91, 1.68) | 1.28 (0.94, 1.74) | 1.22 (0.89, 1.65) |
| 3 | 121/51407 | 1.70 (1.26, 2.29) | 1.55 (1.14, 2.09) | 1.46 (1.08, 1.97) | 1.28 (0.94, 1.75) | 1.41 (1.04, 1.92) | 1.20 (0.87, 1.63) |
| P for trend |  | <0.0001 | 0.005 | 0.016 | 0.130 | 0.030 | 0.297 |
| **Occupational classification** |  |  |  |  |  |  |  |
| Managers, senior officials, etc | 154/76773 | 1.0 (ref) | 1.0 (ref) | 1.0 (ref) | 1.0 (ref) | 1.0 (ref) | 1.0 (ref) |
| Administrative, secretarial, etc | 42/29676 | 0.71 (0.51, 1.01) | 0.71 (0.51, 1.01) | 0.70 (0.50, 0.99) | 0.65 (0.46, 0.91) | 0.64 (0.45, 0.90) | 0.59 (0.42, 0.84) |
| Personal service, sales, etc | 48/21630 | 1.12 (0.81, 1.55) | 1.08 (0.78, 1.50) | 1.04 (0.75, 1.45) | 0.90 (0.65, 1.25) | 0.86 (0.61, 1.21) | 0.75 (0.53, 1.06) |
| P for trend |  | 0.993 | 0.887 | 0.713 | 0.219 | 0.145 | 0.027 |

<sup>1</sup>Comorbidity includes diagnoses of vascular or heart disease, diabetes, chronic bronchitis or emphysema, asthma, and hypertension defined according to measured blood pressure and/or use of anti-hypertensive medication. <sup>2</sup> Lifestyle factors includes body mass index, smoking status, alcohol intake frequency & number of types of physical activity taken in last four weeks. <sup>3</sup> Psychological factors include psychological distress, psychiatric consultation, neuroticism, verbal and numerical reasoning, & reaction time.

**Supplemental Table 3.** Odds ratios (95% CI) for the relation of psychological factors with COVID-19 hospitalisation *– based on complete data*

|  | Case no./<br>Risk no.<sup>1</sup> | Adjustments |  |  |  |  |  |
| --- | --- | --- | --- | --- | --- | --- | --- |
|  |  | Age & sex | Age, sex & ethnicity | Age, sex, ethnicity & comorbidity<sup>1</sup> | Age, sex, ethnicity & lifestyle factors<sup>2</sup> | Age, sex, ethnicity & socioeconomic factors<sup>3</sup> | Adjusted for all covariates |
| **Psychological distress** |  |  |  |  |  |  |  |
| 1 (low) | 149/96723 | 1.0 (ref) | 1.0 (ref) | 1.0 (ref) | 1.0 (ref.) | 1.0 (ref.) | 1.0 (ref) |
| 2 | 161/93337 | 1.14 (0.91, 1.42) | 1.15 (0.92, 1.44) | 1.12 (0.89, 1.40) | 1.09 (0.87, 1.36) | 1.13 (0.91, 1.42) | 1.07 (0.86, 1.34) |
| 3 | 113/555059 | 1.36 (1.06, 1.75) | 1.32 (1.03, 1.70) | 1.25 (0.97, 1.61) | 1.15 (0.89, 1.48) | 1.22 (0.95, 1.57) | 1.09 (0.84, 1.40) |
| P for trend |  | 0.016 | 0.028 | 0.080 | 0.272 | 0.107 | 0.501 |
| Per SD increase |  | 1.18 (1.08, 1.29) | 1.16 (1.06, 1.27) | 1.13 (1.03, 1.24) | 1.09 (1.00, 1.20) | 1.12 (1.02, 1.22) | 1.07 (0.97, 1.17) |
| **Psychiatric consultation** |  |  |  |  |  |  |  |
| No | 412/238768 | 1.0 (ref) | 1.0 (ref) | 1.0 (ref) | 1.0 (ref) | 1.0 (ref) | 1.0 (ref) |
| Yes | 58/26798 | 1.26 (0.96, 1.67) | 1.29 (0.98, 1.70) | 1.26 (0.95, 1.66) | 1.19 (0.90, 1.57) | 1.22 (0.92, 1.61) | 1.15 (0.87, 1.52) |
| **Neuroticism** |  |  |  |  |  |  |  |
| 1 (low) | 119/69458 | 1.0 (ref) | 1.0 (ref) | 1.0 (ref) | 1.0 (ref) | 1.0 (ref) | 1.0 (ref) |
| 2 | 191/109358 | 1.03 (0.82, 1.30) | 1.05 (0.83, 1.32) | 1.03 (0.81, 1.29) | 1.02 (0.81, 1.28) | 1.03 (0.82, 1.29) | 1.01 (0.80, 1.27) |
| 3 | 157/85968 | 1.09 (0.85, 1.39) | 1.11 (0.87, 1.41) | 1.06 (0.83, 1.36) | 1.03 (0.81, 1.32) | 1.06 (0.83, 1.35) | 1.00 (0.79, 1.28) |
| P for trend |  | 0.491 | 0.395 | 0.612 | 0.804 | 0.660 | 0.982 |
| Per SD increase |  | 1.03 (0.94, 1.13) | 1.04 (0.94, 1.14) | 1.02 0.93, 1.12) | 1.00 (0.91, 1.10) | 1.01 (0.92, 1.11) | 0.99 (0.90 1.09) |
| **Verbal numerical reasoning** |  |  |  |  |  |  |  |
| 1 (low) | 81/26374 | 2.52 (1.83, 3.47) | 2.09 (1.50, 2.93) | 2.03 (1.45, 2.84) | 1.86 (1.33, 2.62) | 2.06 (1.44, 2.96) | 1.98 (1.38, 2.84) |
| 2 | 86/41404 | 1.70 (1.24, 2.34) | 1.61 (1.17, 2.21) | 1.59 (1.16, 2.18) | 1.53 (1.11, 2.11) | 1.60 (1.16, 2.21) | 1.58 (1.14, 2.18) |
| 3 | 70/57112 | 1.0 (ref) | 1.0 (ref) | 1.0 (ref) | 1.0 (ref) | 1.0 (ref) | 1.0 (ref) |
| P for trend |  | <0.0001 | <0.0001 | <0.0001 | <0.0001 | <0.001 | <0.0001 |
| Per SD decrease |  | 1.46 (1.27, 1.66) | 1.34 (1.17, 1.54) | 1.32 (1.15, 1.52) | 1.27 (1.10, 1.46) | 1.33 (1.15, 1.55) | 1.31 (1.12, 1.52) |
| **Reaction time** |  |  |  |  |  |  |  |
| 1 (low) | 164/99395 | 1.0 (ref) | 1.0 (ref) | 1.0 (ref) | 1.0 (ref) | 1.0 (ref) | 1.0 (ref) |
| 2 | 155/88713 | 1.09 (0.87 1.36) | 1.05 (0.84, 1.31) | 1.04 (0.84, 1.31) | 1.03 (0.82, 1.29) | 1.03 (0.82, 1.29) | 1.02 (0.81, 1.28) |
| 3 | 148/76914 | 1.21 (0.96, 1.53) | 1.10 (0.87, 1.39) | 1.09 (0.86, 1.37) | 1.05 (0.83, 1.33) | 1.04 (0.82, 1.31) | 1.02 (0.80, 1.29) |
| P for trend |  | 0.104 | 0.424 | 0.496 | 0.659 | 0.759 | 0.869 |
| Per SD increase |  | 1.16 (1.06, 1.27) | 1.10 (1.01, 1.21) | 1.09 (1.00, 1.20) | 1.08 (0.99, 1.19) | 1.07 (0.98, 1.17) | 1.06 (0.97, 1.17) |

<sup>1</sup> Comorbidity includes diagnoses of vascular or heart disease, diabetes, chronic bronchitis or emphysema, asthma, and hypertension defined according to measured blood pressure and/or use of anti-hypertensive drugs. <sup>2</sup> Lifestyle factors included body mass index, smoking status, alcohol intake frequency & number of types of physical activity taken in last four weeks. <sup>3</sup> Socioeconomic factors included occupational classification, highest educational attainment, Townsend deprivation index, & household income before tax

**Supplemental Table 4.** Odds ratios (95% CI) for the relation of socioeconomic factors with COVID-19 hospitalisation *– impact of adjusting for biomarkers*

|  |  | Adjustments |  |  |
| --- | --- | --- | --- | --- |
|  | Case no./Risk no.<sup>1</sup> | Age, sex & ethnicity | Age, sex, ethnicity & biomarker<sup>2</sup> | All covariates<sup>3</sup> |
| **Educational attainment** |  | N=420502 | N=301981 | N=108462 |
| Degree | 229/137717 | 1.0 (ref) | 1.0 (ref) | 1.0 (ref) |
| Other qualifications | 406/214337 | 1.19 (1.01, 1.41) | 1.15 (0.93, 1.41) | 0.94 (0.66, 1.33) |
| No qualifications | 241/70003 | 2.07 (1.71, 2.50) | 1.82 (1.43, 2.31) | 1.10 (0.68, 1.77) |
| P for trend |  | <0.0001 | <0.0001 | 0.802 |
| **Annual household income** |  | N=363175 | N=261825 | N=96562 |
| <£18,000 | 241/81207 | 1.89 (1.51, 2.35) | 1.47 (1.12, 1.93) | 1.03 (0.62, 1.72) |
| £18,000-£30,999 | 179/92461 | 1.27 (1.01, 1.60) | 1.09 (0.83, 1.44) | 1.04 (0.65, 1.68) |
| £31,000-£51,999 | 167/95454 | 1.17 (0.94, 1.47) | 1.10 (0.84, 1.49) | 1.13 (0.72, 1.78) |
| ≥£52,000 | 141/95097 | 1.0 (ref) | 1.0 (ref) | 1.0 (ref) |
| P for trend |  | <0.0001 | 0.007 | 0.988 |
| **Neighbourhood deprivation** |  | N=427986 | N=301418 | N=108898 |
| 1 (low) | 205/143483 | 1.0 (ref) | 1.0 (ref) | 1.0 (ref) |
| 2 | 267/143548 | 1.29 (1.07, 1.55) | 1.14 (0.91, 1.42) | 1.10 (0.76, 1.58) |
| 3 | 436/143517 | 1.97 (1.66, 2.34) | 1.58 (1.28, 1.96) | 0.97 (0.66, 1.42) |
| P for trend |  | <0.0001 | <0.0001 | 0.840 |
| **Occupational classification** |  | N=307262 | N=221854 | N=91388 |
| Managers, senior officials, etc | 324/175637 | 1.0 (ref) | 1.0 (ref) | 1.0 (ref) |
| Administrative, secretarial, etc | 94/74137 | 0.69 (0.55, 0.87) | 0.65 (0.49, 0.87) | 0.48 (0.30, 0.77) |
| Personal service, sales, etc | 149/58915 | 1.30 (1.07, 1.59) | 1.28 (1.01, 1.64) | 0.75 (0.48, 1.15) |
| P for trend |  | 0.091 | 0.223 | 0.054 |

<sup>1</sup> Numbers based on unadjusted model. <sup>2</sup>Biomarkers included FEV1, and blood concentrations of c-reactive protein, HbA1c, and HDL cholesterol. <sup>3</sup>Multivariate model included age, sex, ethnicity, diagnoses of vascular or heart disease, diabetes, chronic bronchitis or emphysema, asthma, hypertension defined according to measured blood pressure and/or use of anti-hypertensive drug, body mass index, smoking status, alcohol intake frequency, number of types of physical activity taken in last four week, psychological distress, psychiatric consultation, reasoning, reaction time, FEV1, and blood concentrations of c-reactive protein, HbA1c and HDL cholesterol.

**Supplemental Table 5.** Odds ratios (95% CI) for the relation of psychological factors with COVID-19 hospitalisation – *impact of adjusting for biomarkers*

|  | Case no./Risk no.<sup>1</sup> | Adjustments |  |  |
| --- | --- | --- | --- | --- |
|  |  | Age, sex & ethnicity | Age, sex, ethnicity & biomarkers<sup>2</sup> | All covariates<sup>3</sup> |
| **Psychological distress** |  | N=383655 | N=273998 | N=179391 |
| 1 (low) | 267/153504 | 1.0 (ref) | 1.0 (ref) | 1.0 (ref) |
| 2 | 291/140200 | 1.29 (1.09, 1.53) | 1.16 (0.94, 1.43) | 1.04 (0.79, 1.36) |
| 3 | 224/91205 | 1.51 (1.26, 1.81) | 1.28 (1.01, 1.61) | 0.98 (0.71, 1.34) |
| P for trend |  | <0.0001 | 0.033 | 0.931 |
| Per SD increase |  | 1.19 (1.12, 1.26) | 1.12 (1.04, 1.22) | 1.04 (0.93, 1.17) |
| **Psychiatric consultation** |  | N=426823 | N=303561 | N=194162 |
| No | 751/379080 | 1.0 (ref) | 1.0 (ref) | 1.0 (ref) |
| Yes | 140/487739 | 1.51 (1.26, 1.81) | 1.44 (1.15, 1.82) | 1.27 (0.91, 1.76) |
| **Neuroticism** |  | N=424212 | N=302071 | N=193565 |
| 1 (low) | 224/106910 | 1.0 (ref) | 1.0 (ref) | 1.0 (ref) |
| 2 | 345/174705 | 1.03 (0.87, 1.22) | 0.98 (0.79, 1.22) | 1.03 (0.78, 1.37) |
| 3 | 319/144092 | 1.21 (1.02, 1.44) | 1.19 (0.96, 1.48) | 1.06 (0.78, 1.42) |
| P for trend |  | 0.023 | 0.088 | 0.725 |
| Per SD increase |  | 1.08 (1.01, 1.16) | 1.08 (0.99, 1.17) | 0.99 (0.88, 1.11) |
| **Verbal numerical reasoning** |  | N=174581 | N=122752 | N=89129 |
| 1 (low) | 152/43988 | 2.31 (1.77, 3.02) | 2.35 (1.68, 3.30) | 2.08 (1.33, 3.28) |
| 2 | 115/58446 | 1.45 (1.10, 1.90) | 1.51 (1.07, 2.13) | 1.67 (1.12, 2.49) |
| 3 | 96/72833 | 1.0 (ref) | 1.0 (ref) | 1.0 (ref) |
| P for trend |  | <0.0001 | <0.0001 | 0.001 |
| Per SD decrease |  | 1.37 (1.23, 1.53) | 1.37 (1.19, 1.58) | 1.33 (1.10, 1.61) |
| **Reaction time** |  | N=424432 | N=302492 | N=193832 |
| 1 (low) | 262/140934 | 1.0 (ref) | 1.0 (ref) | 1.0 (ref) |
| 2 | 274/141575 | 1.00 (0.84, 1.19) | 1.01 (0.81, 1.25) | 1.09 (0.83, 1.42) |
| 3 | 345/143368 | 1.16 (0.98, 1.37) | 1.07 (0.87, 1.33) | 1.05 (0.79, 1.40) |
| P for trend |  | 0.078 | 0.505 | 0.724 |
| Per SD increase |  | 1.07 (1.01, 1.14) | 1.06 (0.98, 1.15) | 1.08 (0.96, 1.21) |

<sup>1</sup> Numbers based on age & sex adjusted model. <sup>2</sup> Biomarkers included FEV1, and blood concentrations of c-reactive protein, HbA1c, and HDL cholesterol. <sup>3</sup> Multivariate model included age, sex, ethnicity, diagnoses of vascular or heart disease, diabetes, chronic bronchitis or emphysema, asthma, hypertension defined according to measured blood pressure and/or use of anti-hypertensive drug, body mass index, smoking status, alcohol intake frequency, number of types of physical activity taken in last four week, occupational classification, educational attainment, Townsend deprivation index, household income before tax, FEV1, and blood concentrations of c-reactive protein, HbA1c and HDL cholesterol.

